# Variants in *STAU2* associate with metformin response in a type 2 diabetes cohort: a pharmacogenomics study using real-world electronic health record data

**DOI:** 10.1101/2020.03.18.20037218

**Authors:** Yanfei Zhang, Ying Hu, Kevin Ho, Dustin N. Hartzel, Vida Abedi, Ramin Zand, Marc S. Williams, Ming Ta M. Lee

## Abstract

Type 2 diabetes mellitus (T2DM) is a major health and economic burden because of the seriousness of the disease and its complications. Improvements in short- and long-term glycemic control is the goal of diabetes treatment. To investigate the longitudinal management of T2DM at Geisinger, we interrogated the electronic health record (EHR) information and identified a T2DM cohort including 125,477 patients using the Electronic Medical Records and Genomics Network (eMERGE) T2DM phenotyping algorithm. We investigated the annual anti-diabetic medication usage and the overall glycemic control using hemoglobin A1c (HbA1c). Metformin remains the most frequently medication despite the availability of the new classes of anti-diabetic medications. Median value of HbA1c decreased to 7% in 2002 and since remained stable, indicating a good glycemic management in Geisinger population. Using metformin as a pilot study, we identified three groups of patients with distinct HbA1c trajectories after metformin treatment. The variabilities in metformin response is mainly explained by the baseline HbA1c. The pharmacogenomic analysis of metformin identified a missense variant rs75740279 (Leu/Val) for STAU2 associated with the metformin response. This strategy can be applied to study other anti-diabetic medications. Such research will facilitate the translational healthcare for better T2DM management.

## Introduction

Diabetes mellitus affects 30.2 million adults in the United States, 90-95% of whom have type 2 diabetes mellitus (T2DM) ^1^. Diagnosed diabetes is a major economic burden with an estimated direct and indirect cost in the United States of $327 billion in 2017 ^2^. Quality of life in T2DM patients is affected by complications which can severely impair a patient’s mobility and vision and can increase the risk of heart and kidney diseases. Improvements in both short- and long-term glycemic control is the goal of diabetes treatment as it can delay disease onset and reduce the severity of T2DM-related outcomes ^3-5^. Although lifestyle modification and weight loss have been shown to delay or even prevent T2DM, antidiabetic drugs targeting different pathophysiological defects of T2DM are indispensable for durable glycemic control ^6,7^. Several classes of antidiabetic drugs are in clinical use, including metformin, sulfonylureas (SU), thiazolidinediones (TZD), and the newer classes of medications, such as the glucagon-like peptide-1 receptor (GLP-1R) agonists, dipeptidyl peptidase 4 (DPP4) inhibitors and the sodium-glucose cotransporter-2 (SGLT2) inhibitors that showed better cardiovascular, renal and weight-loss outcomes ^8,9^.

Metformin is the most frequently used oral anti-diabetic medication because of its excellent efficacy, low cost, weight neutrality, good safety profile, and other benefits including improvements in certain lipids, inflammatory markers, and evidence of cardio protection independent from the drug’s glucose-lowering effect ^10^. It is recommend by the American Diabetes Association, the European Association for the Study of Diabetes, and the American Association of Clinical Endocrinologists as the first-line therapy along with lifestyle modification for the management of T2DM ^9,11^. However, glycemic response to metformin is highly variable at the individual level and the mechanism of action for metformin has not been fully elucidated. Several pharmacogenomics studies on metformin identified variants in *ATM, SLC2A2, CPA6*, and *PRPF31* to be associated with metformin response, which provide some insight into the mechanism of action of metformin ^12-14^. Pharmacogenetic studies on sulfonylureas using candidate gene approach identified polymorphisms in *CYP2C9, KCNJ11, ABCC8, TCF7L2, IRS-1, CDKAL1, CDKN2A/2B, KCNQ1*, and *NOS1AP* genes associated with treatment responses ^15^. These genes affect the pharmacodynamics and pharmacokinetics of SU, the insulin releasing mechanism, the glucose transportation and other mechanisms of T2DM. No GWAS have been performed to study the pharmacogenomics of SU and other newer anti-DM medications. Most of the studies used cross-sectional on-treatment and the baseline Hemoglobin A1c (HbA1c) data. The associations with longitudinal HbA1c responses are not validated. One study has developed a computational model using longitudinal data and performed simulation to evaluate 9 single-nucleotide variants (SNV)s that can predict the long-term HbA1c levels after metformin initiation ^16^.

Advances in documentation and coding in electronic health record (EHR) systems have improved the accuracy of patients’ records on comorbidities, medications, and laboratory tests, thereby serving as a reliable source of real-world clinical data for improving patient care and research. Although the diagnostic criteria for T2DM are well established, the identification of these criteria from EHR data can be challenging ^17^. Several phenotyping algorithms for diabetes have been developed to accelerate research leveraging EHR data ^18^. Among these algorithms, only the eMERGE T2DM algorithm was designed to identify T2DM patients exclusively. It uses information from diagnosis codes, anti-diabetic medications, and laboratory tests, and excludes the type 1 (T1DM) cases ^19^. This phenotyping algorithm can be applied for a wide range of purposes, including genetic studies ^19^. The algorithm showed a very good specificity (0.99) in identifying T2DM patients from EHR data ^18^. As an eMERGE study site, Geisinger has tested and implemented the eMERGE T2DM algorithm on its deidentified EHR database.

In this study, we identified and characterized a T2DM cohort and assessed the impact of pharmacogenomics on disease management of a rural population leveraging the longitudinal EHR data at Geisinger, a single integrated healthcare delivery system. Taking metformin as an example, we identified groups of patients with differing responses and performed a pharmacogenomics genome-wide association study (GWAS) to identify associated genetic variants that could explain, in part, the different responses.

## Cohort and Methods

### Study Cohort

The study cohort included 125,477 patients identified from the Geisinger electronic health records (EHR) data warehouse using the eMERGE T2DM algorithm ^20^. We included 106,190 patients with a current age between 18 to 84 years in the analyses. A subset of these patients also participated in the Geisinger MyCode Community Health Initiative (MyCode®), a system-wide research biorepository at Geisinger with more than >260,000 participants enrolled to date. Participants are consented to use their deidentified genetic and EHR data for research purposes ^21,22^. This study was waived from Institutional Review Board approval because only deidentified data were used. We received MyCode Governing Board approval to perform genetic studies.

### Data extraction

The phenomic analytics and clinical data core at Geisinger applied the eMERGE T2DM algorithm (Supplementary figure 1) to the deidentified data and extracted the data from the inception of the EHR up to July 31, 2018. Demographic information, International Classification of Diseases (ICD)-9 and 10 diagnosis codes, medication prescription, laboratory test results including HbA1c and serum creatinine, and weight and blood pressure at each encounter were retrieved for analysis. Unit harmonization was performed for the quantitative measurements to ensure the high data quality. We estimated the estimated glomerular filtration rate (eGFR) using the CKD-Epidemiology Collaboration equation ^23,24^. All-cause mortality was identified based on the vital status, which is updated biweekly from the Social Security Death Index. Major adverse cardiovascular events (MACE) is defined as a collection of the following clinical events: myocardial infarction, percutaneous coronary intervention, and coronary artery bypass, all of which were identified from EHR by using ICD-9 and ICD-10 codes (Supplementary table 1). A previous study had demonstrated good diagnostic accuracy of this code-based EHR-derived MACE at Geisinger ^25^.

**Table 1.**
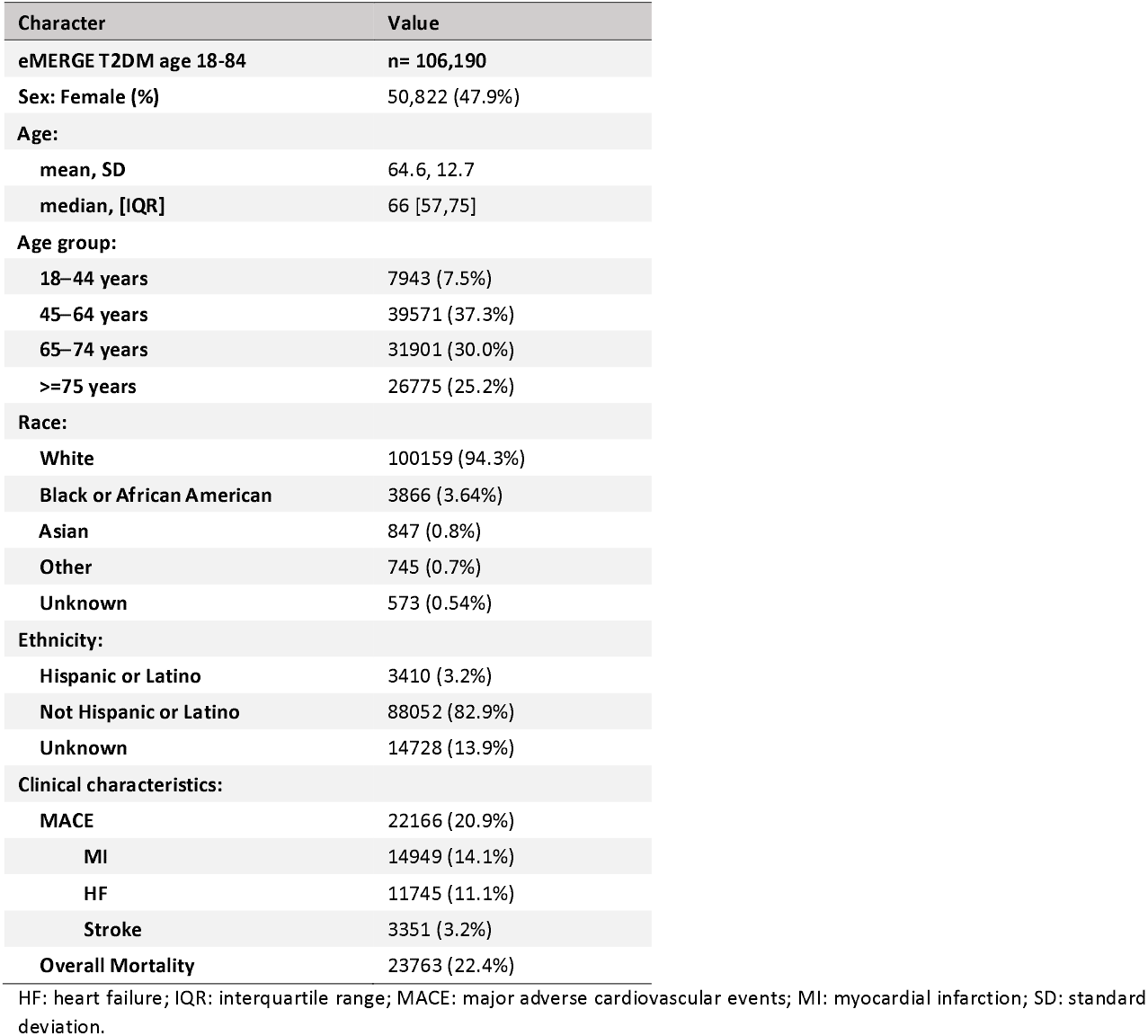
Characterization of the T2DM cohort identified by eMERGE algorithm

| Character | Value |
| --- | --- |
| **eMERGE T2DM age 18-84** | **n= 106,190** |
| **Sex: Female (%)** | 50,822 (47.9%) |
| **Age:** |  |
| mean, SD | 64.6, 12.7 |
| median, [IQR] | 66 [57,75] |
| **Age group:** |  |
| 18–44 years | 7943 (7.5%) |
| 45–64 years | 39571 (37.3%) |
| 65–74 years | 31901 (30.0%) |
| >=75 years | 26775 (25.2%) |
| **Race:** |  |
| White | 100159 (94.3%) |
| Black or African American | 3866 (3.64%) |
| Asian | 847 (0.8%) |
| Other | 745 (0.7%) |
| Unknown | 573 (0.54%) |
| **Ethnicity:** |  |
| Hispanic or Latino | 3410 (3.2%) |
| Not Hispanic or Latino | 88052 (82.9%) |
| Unknown | 14728 (13.9%) |
| **Clinical characteristics:** |  |
| MACE | 22166 (20.9%) |
| MI | 14949 (14.1%) |
| HF | 11745 (11.1%) |
| Stroke | 3351 (3.2%) |
| Overall Mortality | 23763 (22.4%) |

HF: heart failure; IQR: interquartile range; MACE: major adverse cardiovascular events; MI: myocardial infarction; SD: standard deviation.

**Figure 1.**
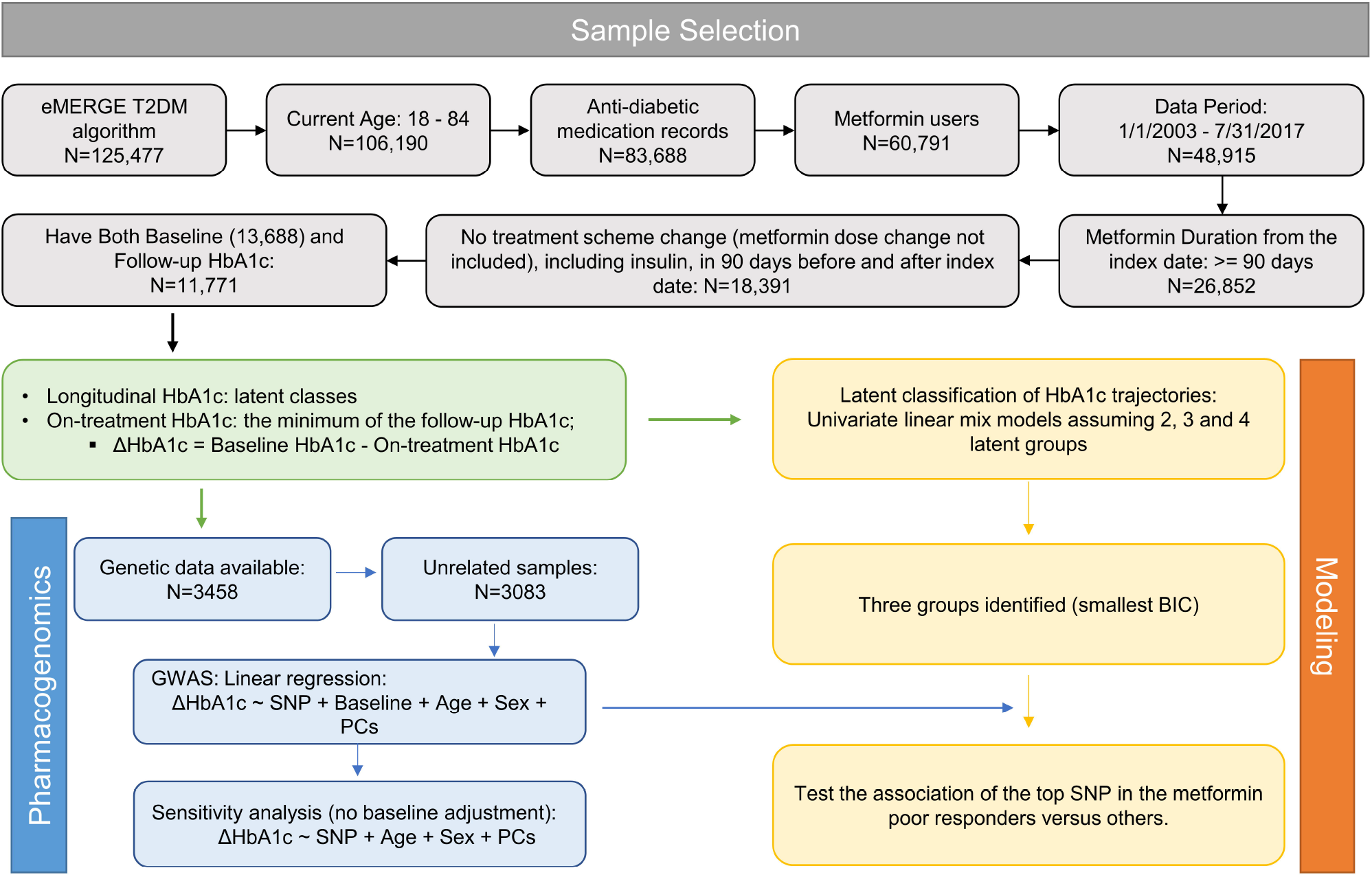
Study design and flowchart. The grey boxes are steps of sample selection; the green box is the summary of the study design; the blue boxes represent the genome-wide association tests and a sensitivity analysis. The yellow boxes represent the linear mixed effect modeling of the longitudinal HbA1c outcome.

### Phenotype definitions for metformin response

We included data from January 1, 2003 to July 31, 2017 to study the glycemic control in patients receiving metformin. We chose these dates in part because the number of new patients on metformin grew steadily after 2003. The index date is defined as the date of the first metformin prescription and must be after January 1, 2003 and before July 31, 2017 to allow a minimum of one year of follow-up. Patients who do not have recorded follow up were also excluded. The duration of metformin use needed to be at least 90 days from the index date. Patients with changes in the treatment scheme during the 90 days before and after the index date were excluded from the study. Treatment scheme change is defined as patients started or stopped an anti-diabetic medication, including insulin, during this time window; metformin dose change is not considered treatment scheme changes. Baseline HbA1c is defined as the HbA1c tests performed between 90 days before and 7 days after the index date. If multiple baseline HbA1c tests were available, the one that was closest to the index date was selected. On-treatment HbA1c is defined as the HbA1c test available from 90 days until 540 days (18 months) after the index date or the time of treatment scheme change, whichever came first ^12^. The minimum on-treatment HbA1c values were used to calculate the decrease of HbA1c for metformin treatment: ΔHbA1c = Baseline HbA1c – minimal on-treatment HbA1c. Figure 1 shows the study design and the flowchart using metformin as an example.

### Statistical analyses

A linear regression model is used to determine the significant variables associated with the decrease of HbA1c using R software (version 3.6.0). A trend test was performed using Stata (IC16.0). We employed the lcmm R package (V3.6.0) to identify groups of patients with distinct HbA1c trajectories after metformin treatment ^26^. We used the lcmm() function to estimate the univariate latent class mixed models of HbA1c (%) with the follow-up times (Days). Two, three and four latent classes were assumed and modeled. Bayesian information criterion (BIC) was used to evaluate model performance. Model with smaller BIC is considered better.

### Genome-wide association study

Genotyping and imputation of MyCode genetic data were described previously ^27^. Briefly, variants with minor allele frequency >1%, missingness <1% were included. One of the pairs of related samples were excluded (PI_HAT >= 0.125 determined by the identity by decent function in plink). Finally, 3,882,700 SNVs and 3083 individuals were included in the association tests. A linear model assuming an additive genetic mode was used to identify associated SNVs with ΔHbA1c, adjusting the significant covariates including age, sex, baseline HbA1c, and 4 principal components (PCs). We also performed a sensitivity analysis without adjusting for baseline HbA1c. The top SNVs were also examined to evaluate the association in the metformin poor responder group versus other groups that were identified in the linear mixed modeling. Plink 1.9 was used for genetic data processing and the association tests ^28^. GTEx portal (https://gtexportal.org/home/) was used to query the eQTL and gene expression.

## Results

### Characterization of the T2DM cohort

Using the eMERGE T2DM algorithm, we identified 106,190 patients with T2DM and current age between 18 to 84 years old (Table 1). Forty-eight percent of the patients are women. Most patients are elderly, only 7.5% of the patients are younger than 45 years old. The majority of our patient population is white (94.3%) and non-Hispanic (82.9%). During the study period, about 20.0% of the patients developed MACE, and the overall mortality was 22.4%.

We then looked at the anti-diabetic medication use and the overall HbA1c values of the T2DM cohort. A total of 83,688 patients (78.8%) had anti-diabetic medication prescription records available; 92,784 patients (87.4%) had at least one HbA1c test value; and 78,913 patients (74.3%) had both medication and HbA1c data. Supplementary table 2 lists the total number of prescriptions and patients on each anti-diabetic medication class. Figure 2A-D shows the number of total patients (A, B) and number of new patients (C, D) on each drug class by year. Metformin (biguanides) and the sulfonylureas are the two most used oral anti-diabetic medications, followed by other newer classes of drugs, such as DPP-4 inhibitors, GLP-1 receptor agonists (GLP-1RA), and the SGLT2 inhibitors. The number of total patients on each drug class increases steadily each year except the TZD (Figure 2B). We observed a system-wide rapid increase in the number of new patients until 2001 and a sudden decrease from 2002-2004 for metformin, sulfonylureas, insulin (Figure 2C) and TZD (Figure 2D). The GLP-1RA, DPP4 inhibitors, and SGLT2 inhibitors emerged in 2005, 2006 and 2013, reflected the time of Food and Drug Administration (FDA) approval of these drugs (Figure 2B and D). The decrease of patient numbers in 2018 is because only 7 months’ data were included. The boxplots of the median HbA1c values of each patient by year were shown in Fig.2E. Median HbA1c values decreased to the clinical glycemic target of 7% in 2002 and fluctuated around 7% ever after. There are extreme HbA1c values over 15% or below 4% every year, indicating large inter-individual variability in glycemic control.

**Table 2:** Characteristics of patients in the metformin study.

|  | Total | GWAS subset | Group 1 | Group 2 | Group 3 |
| --- | --- | --- | --- | --- | --- |
| **N (%)** | 11771 | 3083 (26.2%) | 505 (4.29%) | 10412 (88.45%) | 854 (7.26%) |
| **Age** | 62.5, 12.6 | 63.1, 12.0 | 57.1, 12.8 | 63.0, 12.6 | 58.8, 11.6 |
| **Female sex** | 5568 (47.3%) | 1573 (51.0%) | 211 (41.8%) | 5055 (48.5%) | 302 (35.4%) |
| **Metformin-mono therapy** | 10915 (92.7%) | 2889 (93.7%) | 395 (78.2%) | 9722 (93.4%) | 798 (93.4%) |
| **Baseline HbA1c** | 7.72, 1.60 | 7.62, 1.50 | 9.30, 1.72 | 7.33, 1.06 | 11.60, 1.38 |
| **Minimum HbA1c** | 6.71, 1.18 | 6.62, 1.15 | 10.10, 1.80 | 6.49, 0.76 | 7.32, 1.60 |
| **ΔHbA1c** | 1.01, 1.54 | 0.99, 1.45 | -0.76, 2.15 | 0.83, 1.03 | 4.24, 2.09 |

Categorical features, such as Female sex and metformin-mono therapy, were expressed as number of patients (percentage); Continuous features, such as age, baseline and minimum HbA1c, as well as the ΔHbA1c, were expressed as mean, SD.

**Figure 2.**
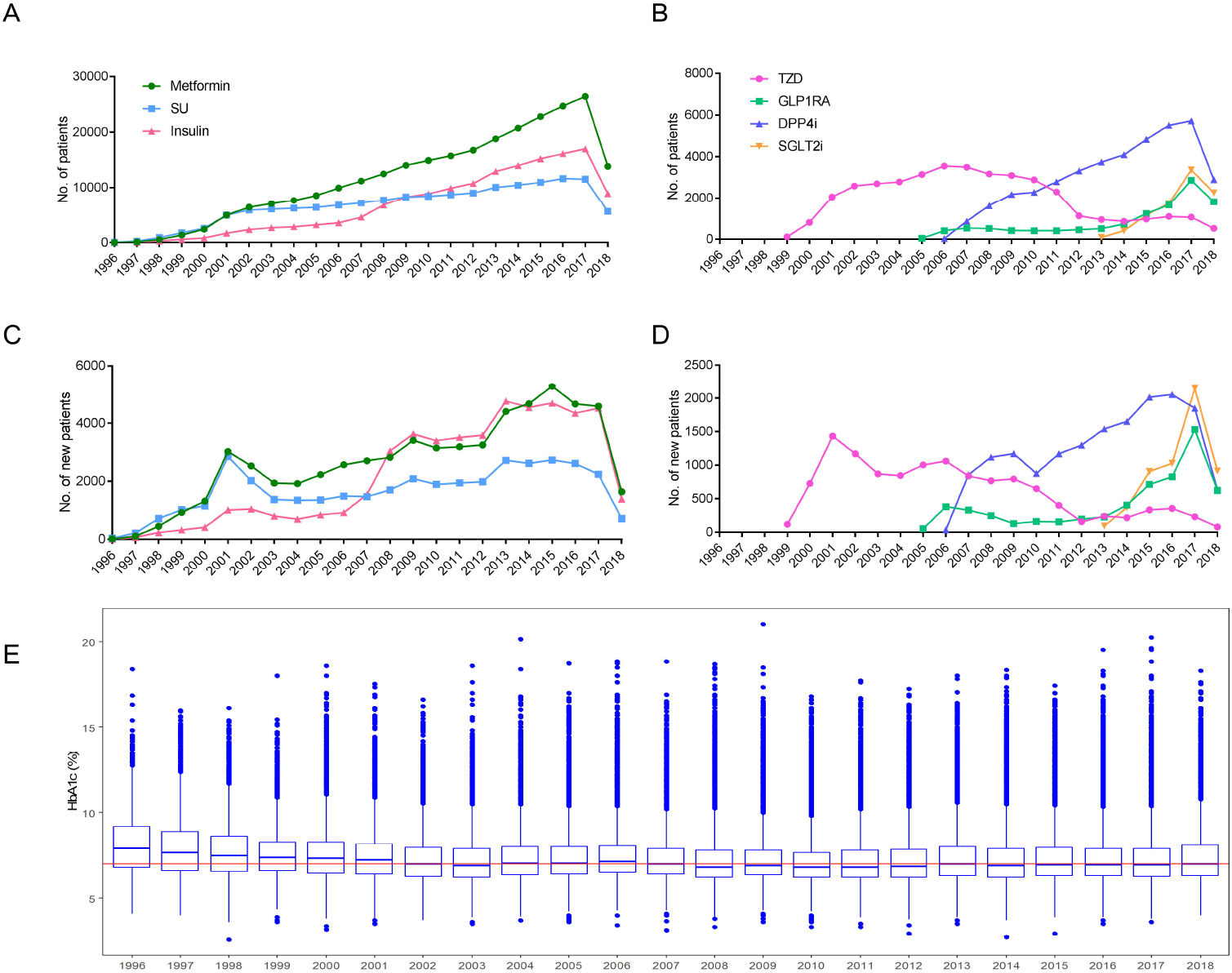
The plot of anti-diabetic medication and the HbA1c of the T2DM cohort by year. A) Total number of patients on metformin (circle), sulfonylurea (SU, square) and insulin (triangle) each year; B) Total number of patients on thiazolidinediones (TZD, circle), GLP1 receptor agonist (GLP1RA, square), DPP4 inhibitors (DPP4i, up-point triangle), and SGLT2 inhibitors (SGLT2i, down-pointing triangle) each year; C) Number of new patients started on metformin, sulfonylurea and insulin each year; B) Number of new patients started on TZD, GLP1 RA, DPP4i, and SGLT2i each year; E) The boxplot of the median HbA1c values of each patient by year. Horizontal line represents HbA1c =7%.

### Variability in Metformin responses

Due to the longest use and the large number of patients available, we selected metformin to evaluate the glycemic control effect. We only evaluated new patients on metformin with an index date from 2003 through July 2017 to allow for at least a one-year follow up to detect changes in HbA1c. Figure 1 illustrates the details of the sample selection process. In total, we identified 11,771 patients eligible for subsequent analyses. Variations in the baseline HbA1c, minimum of on-treatment HbA1c, and the ΔHbA1c were observed (Figure 3A). The mean ΔHbA1c for all the patients on metformin is 1.01% ±1.54% (Table 2). The baseline HbA1c is significantly and positively correlated with the decrease in HbA1c (Supplementary figure 2; p<2×10^−16^) and explained approximately 52% of the variation in metformin response. Most patients were on metformin monotherapy (10,915, 92.7%). Patients on metformin monotherapy had significantly better response than patients on add-on therapy (mean ΔHbA1c of 1.03 vs 0.77, respectively; p=5.03×10^−8^) despite their lower baseline HbA1c (7.67 vs 8.31, respectively; p<2.2 ×10^−16^). Similar results were observed for the 3,083 unrelated patients with genetic data (Supplementary figure 3): the mean ΔHbA1c is 0.99% ± 1.45%; patients on metformin monotherapy (N=2,889) had better response (mean ΔHbA1c of 1.01 vs 0.75, respectively; p=0.01) and lower baseline HbA1c (7.57 vs 8.28, respectively; p=1.86×10^−11^); the index age, baseline HbA1c, and metformin monotherapy are significantly associated with the ΔHbA1c and explained 50.6% of the variability, of which, the baseline HbA1c itself explained 48.7% (Supplementary table 3).

**Table 3:** Lead variants in each locus associated with the ΔHbA1c or the latent classes

| **a.** |  |  |  | **GWAS** |  | **Sensitivity** |  | **Gene** |
| --- | --- | --- | --- | --- | --- | --- | --- | --- |
| **rsID** | **CHR:BP** | **A1/A2** | **MAF (%)** | **Beta [95% CI]** | **P** | **Beta [95% CI]** | **P** |  |
| rs75740279 | 8:74334931 | C/A | 1.23 | -0.65<br>[-0.87, -0.42] | 1.99E-08 | -0.33<br>[-0.65, -0.0063] | 4.57E-02 | *STAU2/STAU2-AS1* |
| rs72736043 | 5:18457292 | G/A | 1.92 | -0.48<br>[-0.67, -0.29] | 5.00E-07 | -0.43<br>[-0.70, -0.17] | 1.40E-03 | / |
| rs75387644 | 3:64892468 | C/A | 1.51 | -0.51<br>[-0.72, -0.30] | 1.58E-06 | -0.54<br>[-0.83, -0.24] | 3.99E-04 | *ADAMTS9-AS2* |
| rs117714057 | 13:62851261 | C/T | 1.13 | -0.58<br>[-0.82, -0.34] | 2.17E-06 | -0.50<br>[-0.84, -0.16] | 4.08E-03 | / |
| rs75899089 | 6:119125847 | T/G | 1.04 | -0.60<br>[-0.85, -0.35] | 2.92E-06 | -0.53<br>[-0.89, -0.17] | 3.81E-03 | / |
| rs56845226 | 10:25637810 | T/C | 1.05 | -0.59<br>[-0.84, -0.34] | 3.20E-06 | -0.45<br>[-0.81, -0.095] | 1.31E-02 | *GPR158* |
| rs76849998 | 11:95779956 | C/T | 1.08 | -0.58<br>[-0.83, -0.33] | 4.09E-06 | -0.53<br>[-0.88, -0.18] | 3.27E-03 | *MAML2* |
| rs144649395 | 7:14900995 | G/C | 1.91 | 0.43<br>[0.25, 0.62] | 4.29E-06 | 0.48<br>[0.22, 0.75] | 3.11E-04 | *DGKB* |
| rs7903977 | 10:99155055 | T/C | 1.93 | -0.43<br>[-0.62, -0.25] | 4.68E-06 | -0.39<br>[-0.65, -0.13] | 3.54E-03 | *RRP12* |
| rs77273237 | 7:6084610 | C/G | 1.06 | -0.58<br>[-0.83, -0.33] | 4.84E-06 | -0.55<br>[-0.91, -0.20] | 2.28E-03 | *EIF2AK1* |
| rs147659285 | 2:164702463 | C/A | 1.28 | -0.53<br>[-0.76, -0.31] | 4.92E-06 | -0.36<br>[-0.69, -0.034] | 3.04E-02 | *AC092684.1* |
| **b.** |  |  |  | **Group** | **Sample size** | **OR [95% CI]** | **P** | **Gene** |
| rs75740279 | 8:74334931 | C/A | 1.23 | Class1 vs. 2&3 | 110 vs. 2973 | 2.43 [1.04, 5.67] | 0.040 | *STAU2/STAU2-AS1* |
|  |  |  |  | Class1 vs. 2 | 110 vs. 2800 | 2.41 [0.98, 5.94] | 0.055 |  |
|  |  |  |  | Class1 vs. 3 | 110 vs. 173 | 1.50 [0.39, 5.84] | 0.557 |  |

CHR: chromosome; BP: base pair; A1: the risk allele also the minor allele; A2: the reference allele; MAF: minor allele frequency; OR: odds ratio.

**Figure 3.**
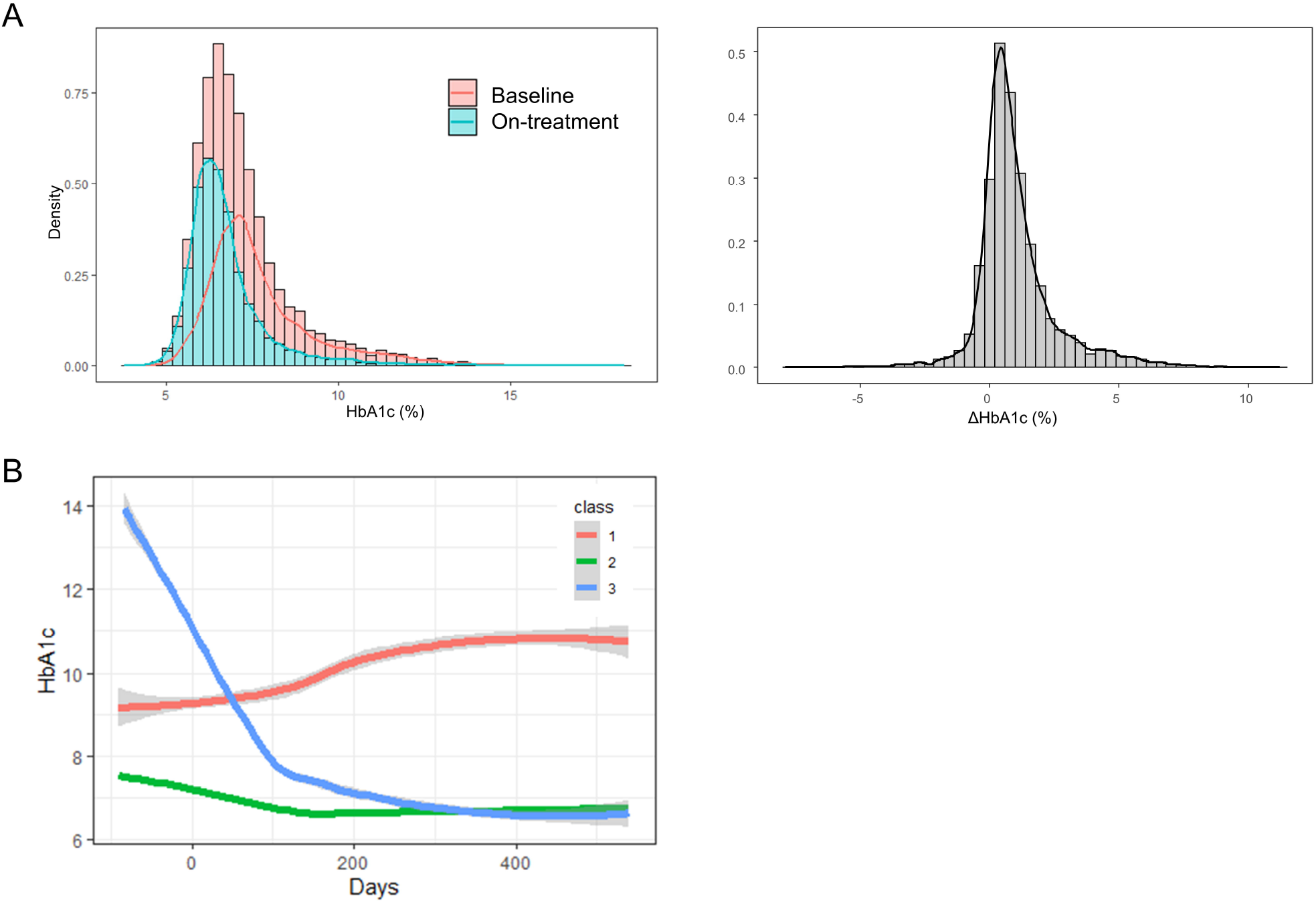
Variabilities of the HbA1c in the metformin treatment. A) Density and Histogram of the Baseline HbA1c (pink), the minimum of the on-treatment HbA1c (cyan), and the ΔHbA1c (right panel, grey); B) The HbA1c trajectories of the three latent classes identified by the lcmm model.

ΔHbA1c is determined by the minimum value of the on-treatment HbA1c, which represents a cross-sectional time point. With the longitudinal HbA1c in the EHR, we can evaluate the longitudinal metformin responses over the course of treatment. We employed a linear mixed model to identify groups of patients with distinct HbA1c responses (trajectories). We examined 2-4 latent classes and determined that the model assuming 3 latent classes had the smallest BIC (Supplementary table 4). The HbA1c trajectories of the three patient groups are shown in Figure 3B and the individual trajectory for each group in supplementary figure 4. Table 2 describes the characteristics of the patients in the three groups. Most patients were classified into group 2 (88.45%) with the lowest baseline HbA1c among the three groups and had good glycemic control. Group 1 patients, constituting 4.29%, showed increased on-treatment HbA1c. Group 1 patients also had the lowest rate of metformin monotherapy (78.2%). Although having the highest baseline HbA1c, Group 3 patients (7.26%) had the best initial response to metformin: the HbA1c decreased significantly in the first three months and slowly decreased to approximately 6.5% after six months (Figure 3B).

**Figure 4.**
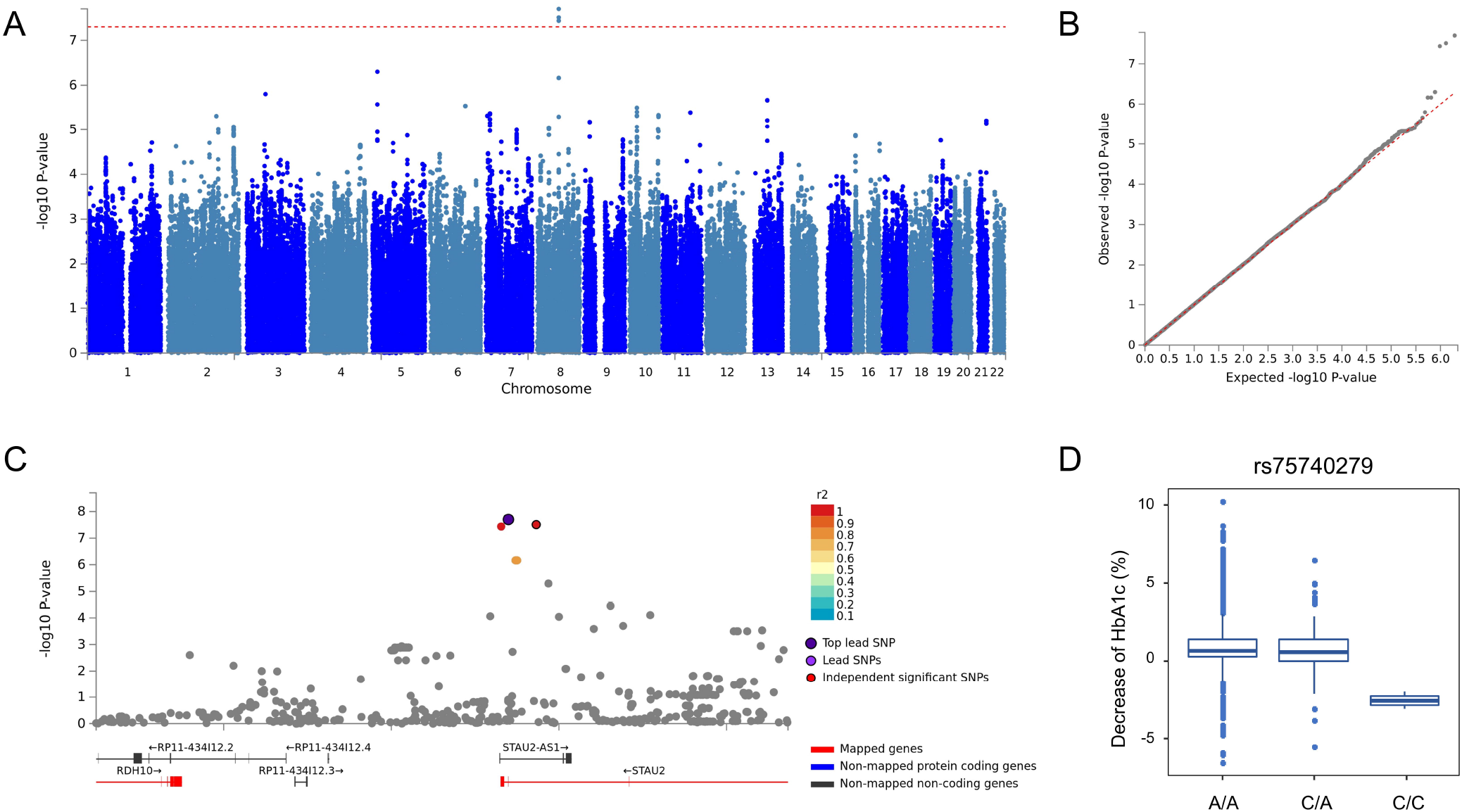
GWAS of the decrease of HbA1c using a linear regression model adjusting for the Baseline HbA1c, monotherapy, PCs. A) Manhattan plot; B) QQ plot; C) Reginal association plot for the genome-wide significant locus XX; D) Boxplot of the decrease of HbA1c by the genotype of rs75740279.

### Pharmacogenomics of the Metformin responses

A GWAS was then performed to identify genetic variants associated with changes in metformin response. Figure 4A-B showed the Manhattan and QQ plot of the GWAS results adjusted for all the significant variables. Table 3a listed the lead SNV in each locus that has associated p value < 5×10^−6^. We identified one genome-wide significant locus on 8q21.11 harboring STAU2 and STAU2-AS1. Figure 4C shows the regional association plot. The lead SNV rs75740279 is a missense SNV (Leu/Val) for STAU2. The minor allele C (allele frequency of 1.23%) is negatively associated with the ΔHbA1c (beta = −0.65, 95% CI [−0.87, −0.42], p=1.99×10^−8^). Figure 3D shows the boxplot of the ΔHbA1c by the genotype of rs75740279 (p_trend_ = 0.043). Two patients with the C/C genotype had the worst response (HbA1c increased by 2.55% after treatment), while the C/A group had smaller ΔHbA1c than the A/A group (0.83 % vs 1%).

In the sensitivity analysis without the baseline HbA1c adjustment, we did not identify any genome-wide significant variants (Supplementary figure 5 for the Manhattan and QQ plots). However, the top variants in the primary GWAS remains nominally significant (p<0.05, Table 3).

Previous GWAS identified significant associations of rs11212617 (11:108283161) in ATM ^12^ and rs8192675 (3:170724883) in SLC2A2 ^13^ with the reduction of HbA1c. Although we did not see significant associations for the 2 SNVs in our study (p=0.28 and 0.52, respectively), we found several SNVs that are independent from these SNVs (the linkage disequilibrium r2 < 0.5) and have marginal association with the decrease of HbA1c (p<0.05, supplementary table 5) with the same direction of effect and larger effect sizes.

We also examined the association of rs75740279 by comparing Group 1 with all others, or with Group 2 or 3. Rs75740279 associated with the Group 1 (poor responders) at a nominal significance level (p<0.05) when compared with all others, with a marginal significance level (p= 0.055) when compared with Group 2 patients alone (Table 3b). However, it is not significant (p=0.557) when compared with Group 3, although these two groups represent two extreme phenotypes.

## Discussion

In this study, we characterized a T2DM cohort in a rural population identified from Geisinger’s EHR database using the eMERGE T2DM algorithm. Leveraging real-world data, we identified a subset of patients with a history of metformin use as a pilot study to evaluate the longitudinal glycemic controls of metformin, and a pharmacogenomic study to identify genetic variants associated with metformin treatment response.

There is increasing concern for diabetes in rural communities, which have 17% higher rates of T2D than urban communities ^29^. Geisinger’s service area includes most of the rural counties in central and northeast Pennsylvania, which are located in region known as diabetes belt ^30^. Despite the association of rurality with increased rate of T2DM, rural populations remain underrepresented in research in general. Thus, there is an unmet need to understand how T2DM is managed in these regions. In this study, we identified 106,910 T2DM patients, representing approximately 7.25% of patients in the entire Geisinger system. This is very similar to the prevalence of T2DM in non-Hispanic Whites (7.4%) reported by the American Diabetes Association (ADA) ^31^. The median HbA1c values decreased and remained steady around the clinical target of 7% after 2002, indicating generally good management of glycemic control. Although rural areas may have higher prevalence of T2DM, the Geisinger covered population showed a similar prevalence as the general population.

By analyzing the prescription data, we found that metformin and sulfonylureas are the two most prescribed drugs and their use increased steadily over the study duration. The Food and Drug Administration (FDA) approved the first use of thiazolidinediones (TZD), GLP-1RA (Byetta), DPP4 inhibitors (Sitagliptin) and SGLT2 inhibitors (Canagliflozin) in 1999, 2005, 2006 and 2013, respectively. This is also observed in our data with a trend in using the newer classes of medications, especially the rapid increase of the GLP-1RA and SGLT2 inhibitors after 2016, reflecting preferential use of these new drugs after several clinical trials showed the lower risk and rates for cardiovascular events and diabetic kidney disease ^32-35^. The use of TZD started to decrease from 2006, likely due to the increased risk for ischemic myocardial events of the TZD ^36^ and the emergence of newer classes of drugs with better outcomes.

We selected metformin for this initial study because of the large sample size and its relatively simple treatment regimen compared with other anti-diabetic agents. In our study, metformin decreased HbA1c by an average of 1.01%, with large variations among the patients. The baseline HbA1c itself explains almost 50% of the variability in the metformin response. Every 1% increase in the baseline HbA1c is associated with an additional 0.7% decrease of the on-treatment HbA1c. Patients on metformin monotherapy had an additional 0.26% decrease in HbA1c compared with patients on combination therapy, although the latter had higher baseline HbA1c (7.67% vs 8.31%). One strength of this study is the modeling of the longitudinal HbA1c data, which allowed us to identify three groups of patients showing distinct HbA1c trajectories. Although most of the patients have good glycemic control on metformin treatment, both the previous study ^16^ and ours identified a group of patients with increased HbA1c trajectory, and a negative association with age. The disease progression continues to get worse with time according the previous study which had longer follow-ups than ours ^16^. Further analyses of the characteristics of the three groups of patients corroborated the clinical findings in the cross-sectional observations. For example, lower proportions of metformin monotherapy were found in the group of patients with poor glycemic control, indicating that this group of patients does not respond well to metformin monotherapy and requires additional medications to achieve the target HbA1c levels. Because T2DM is a multifactorial disease and anti-diabetic medications target different pathogenic pathways ^6,8^, patients who require combination therapy may have more complicated pathogenic disturbances than those who only need metformin monotherapy, suggesting that the complex T2DM pathogenic background also affects the treatment response.

We identified a genome-wide significant locus on 8q21.11 harboring the genes STAU2 and STAU2-AS1 and validated the association of the lead SNV, rs75740279, in the poor responders versus patients in other groups with different HbA1c trajectories. rs75740279 is a missense variant of STAU2 gene and is predicted to be deleterious or probably damaging by SIFT and PolyPhen software and thus potentially functionally significant. The minor allele homozygotes of rs75740279 had worse response than the heterozygotes, suggesting a recessive genetic mode of effect. *STAU2* encodes for Staufen homolog 2, a double-stranded RNA (dsRNA)-binding protein. GTEx data showed that *STAU2* is highly expressed in brain and skeletal muscles ^37^. *Stau2* knockout mice showed significant increased total body fat amount (p=3.68×10^−6^) ^38,39^. STAU2, together with its paralog STAU1, is reported to mediate Staufen (STAU)-mediated mRNA decay (SMD), an important regulatory mechanism of myogenesis and adipogenesis ^40^. SMD targets Krüppel-like factor 2 (KLF2) mRNA, which encodes an anti-adipogenic factor that induces caveolin-1, the main component of caveolae in the plasma membrane ^41^. Interestingly, the insulin-responsive glucose transporter GLUT4 is found in the caveolin-rich fraction, and vesicles containing GLUT4 also contain caveolin, suggesting that caveolae may play an important role in the vesicular transport of GLUT4 ^41^. GLUT4 is primarily expressed in the skeletal muscle and adipose tissue, and the traffic of GLUT4 is a major mechanism for glucose uptake in these cells, suggesting GLUT4 plays an important role in the regulation of glycemic homeostasis ^42,43^. Our study along previous studies suggest a role of *STAU2* in the glucose-lowering mechanism through regulating the transport of GLUT4 by targeting KLF2 and caveolin-1.

This study has several limitations. The patients are ascertained from a single healthcare system that covers the rural area of central and northeast Pennsylvania, and the cohort is predominantly composed of white, non-Hispanics of Northern European descent. The results from this study may not apply to populations of other healthcare systems or ethnicities, as the rate of diabetes and the treatment response are geographically variable. However, our methodology can be applied to other cohorts to identify high-risk subjects with poor metformin response. The same strategies can be applied to study other anti-diabetic drugs for glycemic control and determine evidence of cardiovascular and renal benefits in a real-world setting. Small numbers of patients from our system precluded this analysis in this cohort, although as the number of patients with genotype data increases, these studies may be possible. Second, missing data is an unfortunate consequence of using real-world EHR data. Although we have set the study time to reduce bias and applied the eMERGE algorithm that uses multi-modal data, the incomplete data may still lead to an underestimate of the true prevalence of T2DM and the medication adherence evaluation in our population. While the poor responses of some patients might be due to suboptimal medication adherence, the strong and sustained responses observed in groups 2 and 3 argues against this as a significant factor. With future access to the patients’ claims data, we can evaluate adherence using the surrogate measure of prescription fills and refills^44^. Nonetheless, the nature of the big-data approach is likely balanced by the recorded real-life longitudinal EHR data and the identified patients may still represent the T2DM population.

In summary, we identified three groups of patients with distinct HbA1c trajectories after metformin treatment. Patients on other add-on medication with high baseline HbA1c are prone to have worse HbA1c outcome than others. We identified a genome-wide significant missense variant rs75740279 in *STAU2* that is associated with poor response to metformin treatment. The methodology can be applied to study other anti-diabetic drugs. The results need to be validated in other cohorts.

## Data Availability

Please contact the corresponding author for data request.

## Acknowledgements

The authors thank the staff and participants of the MyCode for the integrative work, the staff at Geisinger Phenomic Analytics and Clinical Data Core for EHR database maintenance, and the staff at Regeneron Genetic Center for genetic data processing and support.

## Acronyms

T1DM: type 1 diabetes mellitus
T2DM: type 2 diabetes mellitus
SU: sulfonylureas
TZD: thiazolidinediones
GLP-1R: glucagon-like peptide-1 receptor
DPP4: dipeptidyl peptidase 4
SGLT2: sodium-glucose cotransporter-2
MACE: Major adverse cardiovascular events
EHR: electronic health record
SNVs: single nucleotide variants
GWAS: genome-wide association study

